# The Safety Action Feedback and Engagement (SAFE) Loop: Initial Testing and Refinement of a Novel Intervention to Enhance Hospital Incident Reporting and Patient Safety

**DOI:** 10.1101/2025.06.03.25328744

**Authors:** Edward Seferian, Carl T. Berdahl, Bernice Coleman, Donna Leang, Tara Cohen, Nabeel Qureshi, Sara G. McCleskey, Karen Kaiser, Matthew Grissinger, Falisha Kanji, Andrew J. Henreid, Johan Carrascoza-Bolanos, Laura Daniels, Oscar Abarca, Pamela De La Cruz, Brandon T. Truong, Teryl K. Nuckols

## Abstract

**Background:** Voluntary incident reporting has improved safety in many high-risk industries, but several barriers have limited its effectiveness in hospitals. Designed to overcome these barriers, the novel Safety Action Feedback and Engagement (SAFE) Loop: (1) obtains input from nurses about patient safety problems; (2) invites nursing units to select a Target Event to focus on; (3) teaches nurses to write more informative incident reports; (4) prompts nurses to report Target Events for a designated period; (5) standardizes investigative procedures to support mitigation plans; and (6) provides feedback to nurses about contributing factors and mitigation plans. We aimed to refine the SAFE Loop through iterative testing.

**Methods:** This study reflected Stage I of behavioral intervention development. At a large U.S. academic hospital, we first conducted proof-of-concept testing on two nursing units. Leveraging these experiences and stakeholder input, we made iterative refinements to intervention design, implementation, and evaluation plans. Finally, we conducted structured pilot testing of the refined intervention on one nursing unit. While SAFE Loop could be applied to any patient safety problem, we focused on medication safety.

**Results:** In initial proof-of-concept testing, implementation was feasible and nurses voiced substantial enthusiasm. Following the refinements, the structured pilot test reaffirmed SAFE Loop feasibility, including selecting a Target Event, training nurses in enhanced reporting methods, conducting investigative interviews, developing mitigation plans with nursing unit leaders, and distributing mitigation plans on the unit. The pilot also demonstrated the feasibility of extracting contributing factors from incident reports and surveying nurses about reporting and safety. Qualitative interviews after the pilot reaffirmed intervention acceptability.

**Conclusions:** Nurses found the SAFE Loop to be a promising strategy for improving patient safety. Testing and refinements to the SAFE Loop laid the groundwork for efficacy testing via an ongoing pragmatic randomized controlled trial.

## Introduction

Medical errors continue to harm hundreds of thousands of hospitalized patients in the U.S. annually, and voluntary incident reporting is a widely discussed strategy for improving patient safety. Decades of effort by hospitals and policymakers have improved patient safety in major areas such as healthcare-associated infections [1]. Nonetheless, preventable harms remain common [2, 3]. A 2018 study at 11 hospitals in Massachusetts found that preventable adverse events occurred during 7% of hospitalizations, and 1% of hospitalizations involved serious to fatal events [4]. Earlier studies reached similar conclusions [1–3, 5–8].

Voluntary incident reporting is a technique through which frontline personnel describe events that pose risks, particularly near misses. Incident reporting has two foundational premises. First, a major safety event that causes harm is usually preceded by numerous near misses [9, 10], and near misses and harmful incidents share the same causes [11]. Second, first-hand witnesses can uniquely describe the specific conditions leading up to and potentially causing each incident—insights that might otherwise be rapidly forgotten [12]. By analyzing detailed incident report narratives and conducting follow-up investigations, organizations in high-risk industries can learn how specific work-system factors (e.g., breakdowns associated with technology, the environment, teamwork, task factors or the organization’s culture or resource management) contributed to incidents, directly or by influencing human performance. These insights enable organizations to identify and modify work systems to reduce the chance of events that cause harm. Incident reporting systems with these characteristics have a long history of successful use in the aviation, maritime, chemical, nuclear, and railway industries [13, 14].

Since 1965, incident reporting systems have also been widely used in U.S. hospitals to improve safety and alert risk managers of events that could lead to litigation [15]. Nurses see reporting as a professional responsibility and file most incident reports [16], but they experience barriers to reporting, particularly a lack of training, engagement, and feedback [17, 18]. Nurses are often uncertain about which events to report and what information to include [19]. They fear triggering punitive responses and doubt whether actions will be taken to improve systems of care [19–23]. Although hospitals typically receive thousands of reports annually [24, 25], many reports describe similar events and address lower-risk problems [16]. Higher-risk incidents are underreported [16, 26]. Additionally, only 32-65% of reports reveal actionable insights into the work-system factors that may constitute safety threats [27–30].

The optimal procedures for following up on hospital incident reports have not been described, and how hospitals handle reported incidents varies. Reports may be transmitted to managers in nursing units, hospital safety leaders, and/or risk managers, and they can trigger investigations locally or systemwide. Sometimes reporting leads to discipline of the individuals involved [20, 31–33], and feedback to reporters is rare [18, 22, 34–36]. Hospitals have emphasized the passive collection and classification of large numbers of reports, unlike in other industries, where systematic investigation, prompt action, and feedback to front-line personnel are goals [37–41].

To address these challenges, we designed the Safety Action Feedback and Engagement (SAFE) Loop, a novel intervention designed to transform a hospital’s existing incident reporting and follow-up systems into more effective tools for improving patient safety (as described below). The present work sought to refine the nascent SAFE Loop intervention through preliminary testing. While the SAFE Loop could apply to any patient safety problem, we focused on medication errors because medication administration is a core daily responsibility for nurses, medication errors are common on diverse types of nursing units [42–45], and nurses can detect problems in all stages of medication therapy (ordering, dispensing, administering, and monitoring) [46].

## Materials and Methods

### Overview

Experts recognize several stages in development, testing, and dissemination for interventions designed to shape human behavior in real-world clinical settings. Stage I involves generating and refining a novel intervention via an iterative approach that allows the intervention to remain fluid, involves initial uncontrolled tests with small numbers of selected participants, and permits ongoing refinements in response to evolving findings. Later stages in intervention development involve fully powered experimental testing to establish internal and external validity [47–50].

To generate a novel intervention, developers often start by considering scientific principles, engaging stakeholders, and weighing possible components, and then bringing these together to define key attributes, as we did for the SAFE Loop (below). Next, experts recognize three initial steps in iterative testing and refinement for a nascent intervention:

<u>(A) Proof-of-concept testing</u> occurs in small select samples, using a mixture of qualitative and quantitative data, to shed initial light on the intervention’s feasibility and acceptability to stakeholders;
<u>(B) Iterative refinement of intervention design and implementation plans</u> enables developers to specify details of intervention components and prepare for full implementation; and
<u>(C) Structured pilot testing</u> of the refined intervention yields additional data about feasibility, acceptability, and potential effectiveness [47, 48].

This paper reports steps (A) through (C) for the SAFE Loop intervention. We conducted these steps in preparation for a subsequent pragmatic trial that randomized 20 acute care nursing units to the SAFE Loop intervention versus usual care [51, 52].

### Setting

The work was conducted at a 915-bed major academic hospital in Los Angeles that is staffed by 1,980 nurses and consistently receives Magnet award recognition for excellence in nursing. The hospital employs several systems for improving patient safety. Relevant to medication safety, the hospital has a fully electronic medical record system with clinical decision support, intravenous infusion pumps with clinical decision support, Pyxis^TM^ automated medication dispensing machines (“Pyxis^TM^”), and barcoding systems, among others.

The hospital uses RL6: Risk (RL Solutions, Cambridge, Massachusetts), a risk management and incident reporting software package, for the reporting, classification, root cause analysis, tracking and trending patient safety events, and provision of feedback to leadership with expectations of dissemination to reporters. At baseline, when an incident report is filed from a nursing unit, the nursing unit leadership conducts a brief follow-up review that may include speaking with the reporter and other involved parties. Additionally, the medication safety program team reviews all incidents, has a group of clinicians review each incident to discuss risks and assign levels of harm, conducts more detailed investigations and plans specific actions around common or high-risk events, and summarizes events and mitigation activities annually.

### Participants

Study participants included nursing unit leaders and front-line nurses on three nursing units engaged in SAFE Loop development and testing.

A multidisciplinary research team oversaw development and testing. A physician researcher and a PhD-trained human factors engineer had experience studying patient safety and incident reporting (TN, TC) [16, 27–29, 32]. The team included the Chief Patient Safety Officer (ES), the Director of Nursing Research and Quality (BC), and the Pharmacist Manager for Medication Safety (DL). An external Advisory Board of national leaders in patient safety, medication errors, incident reporting, and nursing practice (see Acknowledgements) provided feedback on refinements in Step (B). Several research methodologists (NQ, KK, XZ) contributed to evaluation activities.

#### Human Subjects/Ethics Disclosure

The Cedars-Sinai Institutional Review Board (IRB) reviewed and approved the current work in two stages. For the first stage, proof-of-concept testing (Pro00055431), the IRB determined that this was institutional quality improvement and not regulated as research. Individually identifiable data were not involved and consent was not obtained. For the second stage, iterative refinement and pilot testing (STUDY00001025), the IRB determining that the SAFE Loop intervention met criteria for a quality improvement activity whereas formal data collection activities constituted research. Written informed consent was obtained for nurse surveys and interviews. Consent was waived for reuse of existing data.

### Key Attributes of the SAFE Loop

To generate ideas about improving hospital incident reporting systems, the study team considered published literature on the systems’ current limitations, including our own prior research [16, 27–29, 32, 53]. In considering strategies for overcoming these limitations, the research team drew from literature on effective strategies for improving the clinical quality of care [54–59], literature on how stakeholder input and engagement can promote the adoption and use of novel interventions [60–63], and the team’s professional experience and knowledge of the institution’s systems and practices.

On this basis, we proposed the SAFE Loop intervention. The SAFE Loop activities are organized around a “Target Event,” meaning a specific medication safety event that nursing unit leaders choose after conferring with front-line nurses. The purpose of the Target Event is to bring a nursing unit together in deciding what medication safety events are particularly problematic, which may focus and promote greater engagement in reporting and subsequent follow-up and mitigation activities. Accordingly, the SAFE Loop has six key attributes (**Table 1**): (1) obtaining input from nurses on the unit about which safety problems to prioritize; (2) focusing on one Target Event safety problem that nursing units select; (3) teaching the nurses to write more informative incident reports; (4) prompting the nurses to report incidents relevant to the Target Event for a designated period; (5) establishing standardized investigative procedures to support development of a mitigation plan; and (6) providing feedback to the nurses about factors contributing to the Target Event and mitigation plan. The attributes occur during three phases: Engagement, Fact-finding, Action and Feedback (see **Figure 1**). These attributes build on, not replace, the existing reporting system.

**Figure 1:**
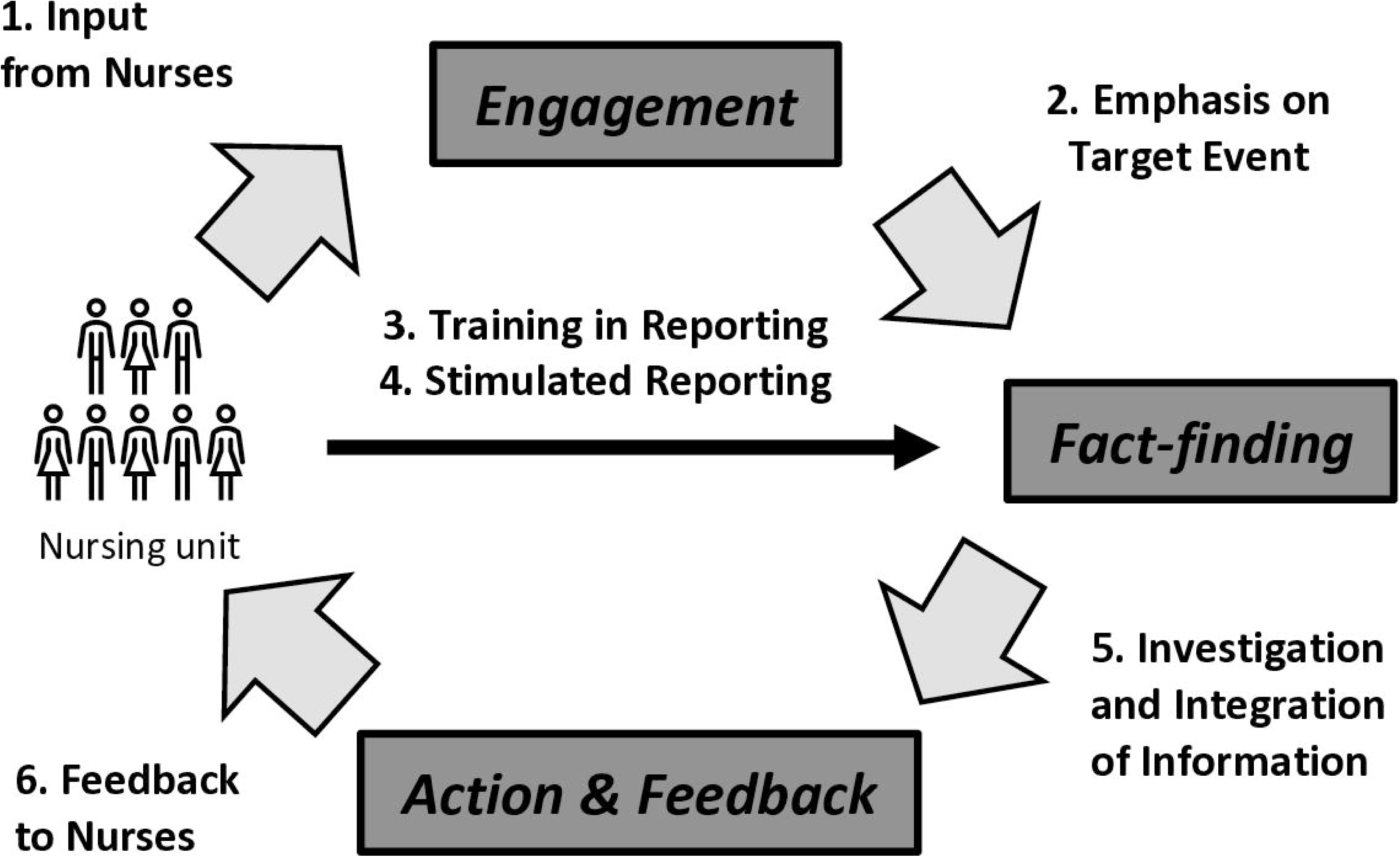
Safety Action Feedback and Engagement (SAFE) Loop with Six Key Attributes Designed to Maximize the Effectiveness of Reporting at Improving Patient Safety.]

**Table 1:** Limitations of Current Hospital Incident Reporting Practices, Conceptual Rationale, and Attributes of the Safety Action Feedback and Engagement (SAFE) Loop.

| Limitations of Current Reporting Practices | Conceptual Rationale | SAFE Loop Key Attributes and Definitions |
| --- | --- | --- |
| Lack of engagement by nurses and doubt that reports are used to improve safety deters reporting [20-22, 34]. | Nurses are more likely to report events and adopt changes for problems that they find to be particularly common, severe, or preventable and/or impactful to the quality and efficiency of nursing practice.[60-62] Engagement is needed between nurses and safety leaders throughout the process of reporting, investigation, and improvement. | **1. Input from Nurses:** The SAFE Loop team solicits input from unit leaders and front-line nurses on which patient safety problem to prioritize for reporting, investigation, and mitigation. |
| Hospitals passively gather thousands of reports on low-priority events each year, making it hard to identify important threats that warrant follow-up and action [16, 24, 25]. | Emphasizing one specific problem with a narrow scope may enable safety leaders to make the best use of investigative resources and obtain enough reports to understand the problem. Common, preventable, and potentially severe events should be a high priority. | **2. Emphasis on a Selected “Target Event”:** The SAFE Loop focuses on each nursing unit's unique *Target Medication Event*. These are errors and adverse events related to a selected, defined problem involving a particular drug class (e.g., opioids), population (e.g., kidney disease), type of error (e.g., duplicate therapy), and/or work-system factor (e.g., conflicting protocols). |
| Lack of guidance about what and how to report deters reporting and limits reports' usefulness [20-22, 34].<br>Gathering rich qualitative insights into contributing factors is central to successful incident reporting but hospital reports provide little detail on contributing factors [9-11, 15, 20-22, 24, 25, 29, 31, 33, 34].<br>When reporting events involving actual harm, nurses are often concerned about being blamed or disciplined. Reports that address harm usually contain little information about contributing work-system factors [28]. | If nurses receive guidance on what and how to report, they would be more likely to submit reports.<br>Report narratives that include more details about contributing work-system factors may be more informative and actionable for efforts to redesign systems and improve safety. A mnemonic may help nurses to remember which types of details to include in incident reports.<br>If reporting emphasizes near misses, this may reduce the risk that reporters might be blamed for the event. | **3. Training in Enhanced Reporting:** The SAFE Loop involves teaching nurses to write more informative incident reports that include details about contributing work-system factors. The SAFE Loop teaches nurses to organize the narrative using the Situation, Background, Assessment, Recommendations (SBAR) format. |
| Hospital incident reporting systems typically function as passive receptacles for incident reports.<br>Nurses are often concerned about the time burden of reporting [20-22, 34]. | Obtaining multiple related reports may provide a more thorough understanding of work-system factors.<br>Asking each nurse to submit one report is a modest request.<br>Having a deadline facilitates progress on a project that involves distinct phases. | **4. Stimulated Time-limited Reporting:** The SAFE Loop prompts nurses to report at least one Target Medication Event, if possible, during a designated period (in addition to any reports that nurses would usually submit). |
| There are no established procedures for conducting follow-up investigations of reported incidents [37, 38].<br>Few safety problems are unique to a particular nursing unit or hospital.<br>Certain AHRQ Patient Safety Organizations systematically examine thousands of incident reports and generate syntheses of their findings. | When interviewed, front-line nurses and other relevant parties may share additional insights about work-system factors.<br>Leveraging publicly available information on the causes of similar events at other hospitals and solutions that have been tested at other hospitals would reduce the work involved and enhance the success of investigative activities and mitigation plans. | **5. Investigation and Integration of Information:** The SAFE Loop integrates information from incident reports, follow-up investigations, interviews with front-line nurses and other relevant parties, and internal and external sources (including published literature and AHRQ Patient Safety Organizations). Standard procedures, including mini-Root Cause Analyses and Ishikawa diagrams, can facilitate investigation and integration and help design a mitigation plan. |
| Nurses seldom receive feedback after filing a report, and a lack of feedback is a major barrier to reporting [20-22, 34]. | If nurses receive feedback and observe improvements in systems of care in response to incident reports, they may have more favorable perceptions of reporting, creating a positive feedback loop that promotes safer care [34]. | **6. Feedback to Nurses:** The SAFE Loop Team provides feedback about the causes of safety problems and mitigation plans. |

### Step (A) Proof-of-concept Testing

Incorporating these key attributes of the nascent SAFE Loop intervention, the Chief Patient Safety Officer oversaw a preliminary test on two nursing units from January to March 2019. One was a neurosurgery/spine floor unit with 78 nurses, and the other was an adult medicine unit with 103 nurses. We selected these units based on having medical-surgical patient populations, engaged nursing staff, and an interested nurse leader who would serve as the intervention champion. Proof-of-concept testing occurred as part of normal operational quality improvement.

#### Intervention

First, we met with unit leaders and nursing staff in each nursing unit to prioritize a patient safety Target Event based on staff nurses’ perceptions of the potential threat and their ability to effect change. Because nurses have limited time to add new responsibilities, we incorporated SAFE Loop implementation into routine daily nursing “huddles” where nurses hear announcements and briefly discuss key issues on the unit; “huddles” occur at the 7 am and 7 pm shift changes. Next, the safety leadership team encouraged nurses to submit examples of the Target Event and emphasized the reporting of near misses. We then collected incident reports about the Target Events occurring on the pilot nursing units. We met weekly with unit leaders and staff during the “huddles” to review the incident reports from each unit. We analyzed the reports and conducted mini-root cause analyses (i.e., root-cause analyses of abbreviated scope) [64]. We met weekly with unit managers and nursing staff to discuss findings from the incident reports and mini-root-cause analyses. Finally, we worked with unit managers to consider potential system changes, obtain feedback, implement tests of change to rectify work-system issues, and rapidly disseminate information.

#### Evaluation

From February 5 through March 25, 2019, we examined the number of incident reports submitted per week on the pilot nursing units as well as the proportions of reports that were near misses (National Coordinating Council for Medication Error Reporting [NCC MERP] categories A and B) or involved a potential for or actual harm (NCC MERP categories C-I) [65, 66]. Additionally, before and after implementation, we surveyed nurses about the culture of safety in the nursing units. The surveys included three items (scale 1-5, 4 = agree, 5 = strongly agree): (1) My unit is assessing and refining processes to improve patient safety before events occur, (2) Staff in my unit do not feel like their mistakes are held against them, (3) Staff in my unit will freely speak up if they see something that may negatively affect patient care (the latter two items were adapted from the AHRQ Hospital Survey on Patient Safety) [67–69]. The culture of safety composite reflected the percentage of positive responses (agree or strongly agree) across the three items. Finally, we asked nursing unit leaders and front-line nurses open-ended questions to obtain feedback about the intervention.

### Step (B) Iterative Refinement of Intervention Design and Implementation

Building on the proof-of-concept testing, we refined and operationalized the SAFE Loop and specified implementation procedures—both during the development of the funding application and after funding. We broke the key attributes down into component steps, proposed a timeline for each step, defined roles in the SAFE Loop and associated tasks (**Table 2**), obtained ongoing input and feedback from safety experts and nursing stakeholders, and established how the team would engage unit directors and personnel on the nursing unit.

**Table 2:** Roles of Personnel in the Existing Reporting System and the SAFE Loop Intervention.

|  | **Position** | **Roles in Existing Reporting System** | **ADDITIONAL Roles in SAFE Loop Intervention** |
| --- | --- | --- | --- |
|  | ***Nurses on Study Nursing Units*** | Spontaneously submit incident reports. | Provide input on the Target Events, undergo training in reporting, submit reports, receive feedback, and implement mitigation strategies. |
|  | ***Nursing Unit Leaders*** | Administratively responsible for overseeing efforts to improve the safety of care on each nursing unit. Participate in analyzing reports, conduct any investigations, design and implement any solutions. | Select the Target Events and work with the SAFE Loop Team throughout implementation. |
| ***SAFE Loop Team*** | ***Incident Report Clinical Reviewer*** | Classifies events, assigns harm severity, identifies source nursing unit, and triages high-risk events to relevant departmental and unit leaders. |  |
|  | ***Patient Safety Leaders (Hospital, Pharmacy, Nursing)*** | Assist nursing units with investigations, obtain resources, engage leaders of relevant departments, and overcome barriers. Oversee root-cause analyses of harmful events. | Part of the SAFE Loop team. Facilitate investigation of Target Events. |
|  | ***Target Event Champion*** | Not applicable. | Obtains input from and trains nurses, identifies and extracts information from reports on Target Events, assists with investigations, integrates information from reports and other sources, drafts feedback to nurses. |
|  | ***AHRQ Patient Safety Organization*** | Not applicable. | Serves as a resource to the SAFE Loop Team, providing information on events at other hospitals. |

Further iterative refinements were continuously made throughout the process of preparing for and conducting the structured pilot testing; we report refinements and implementation under Step (B) and evaluation methods under Step (C), even though in reality, these steps were intertwined.

Refinement and structured pilot testing occurred from March 15, 2022 to November 21, 2023, in partnership with a nursing unit where registered nurses cared for adult patients with stroke and other neurological conditions. We selected this nursing unit because the leaders were enthusiastic about participating and because excluding the unit from the subsequent trial would not affect randomization. During the pilot testing, a reorganization of the unit increased the number of full-time nurses from 26 to 59.

#### Obtaining Input from Nurses and Selecting a Target Event

For the pilot nursing unit, we selected a SAFE Loop “champion,” meaning an individual (generally a clinician) who promoted nurses’ engagement with the SAFE Loop and supported investigative follow-up procedures. The research team and champion met with the nursing unit leadership and then subsequently visited nursing unit huddles to invite nurses to introduce the SAFE Loop and suggest ideas for potential Target Events.

Choosing the Target Event was an iterative process that involved discussions between the research team, champion, and nursing unit leaders to balance multiple considerations. These included nursing leaders’ and front-line nurses’ perceptions of frequency, risk, and persistence despite prior efforts to address the issues; the ability to clearly and precisely delineate the event in a manner that nurses would recognize; and the likelihood that a few specific actions could be taken to mitigate the event, among others. Once the Target Event was selected, we revisited the huddles to inform the nurses and encourage them to submit incident reports about the Target Event over three months.

#### Training in Enhanced Reporting

Our team’s nurse researcher and human factors expert created an online training program to introduce nurses to the SAFE Loop and demonstrate how to write more informative incident report narratives. As reported previously [70], the online training emphasized four key messages for nurses to: (1) participate in the selection and reporting of a high-priority Target Medication Event for their nursing unit; (2) emphasize the reporting of near misses over events involving harm; (3) describe in detail any contributing work-system factors in the narrative portion of the reports on Target Events, and (4) use the Situation, Background, Assessment, Recommendation (SBAR) format to organize their incident report narratives [71]. The training explained how work-system factors contribute to safety events, and defined five broad categories of factors (people, tools/technology, task, environment, and organization), drawn from the Systems Engineering Initiative for Patient Safety (SEIPS) model.[72] The training program incorporated the SBAR format because it is widely used in quality improvement as a framework for communication among healthcare team members. SBAR is an easy-to-remember, concrete mechanism that can frame critical communications about issues that require urgent attention and action, facilitating teamwork in patient safety situations [71]. We considered work-system factors to be an aspect of the Background. To engage nurses, the training program included quizzes about work-system factors. It also provided two in-depth hypothetical examples that compared “typical” versus “enhanced” incident port narratives that used the SBAR format and described work-system factors.

After input and feedback from the core research team, we conducted cognitive tests with four front-line nurses and refined our training. We programmed the novel training using EasyGenerator (Rotterdam, The Netherlands). We also created “badge buddies” (plastic cards worn under the ID badge for easy access) that summarized key messages from the training (Table 3).

**Table 3:**
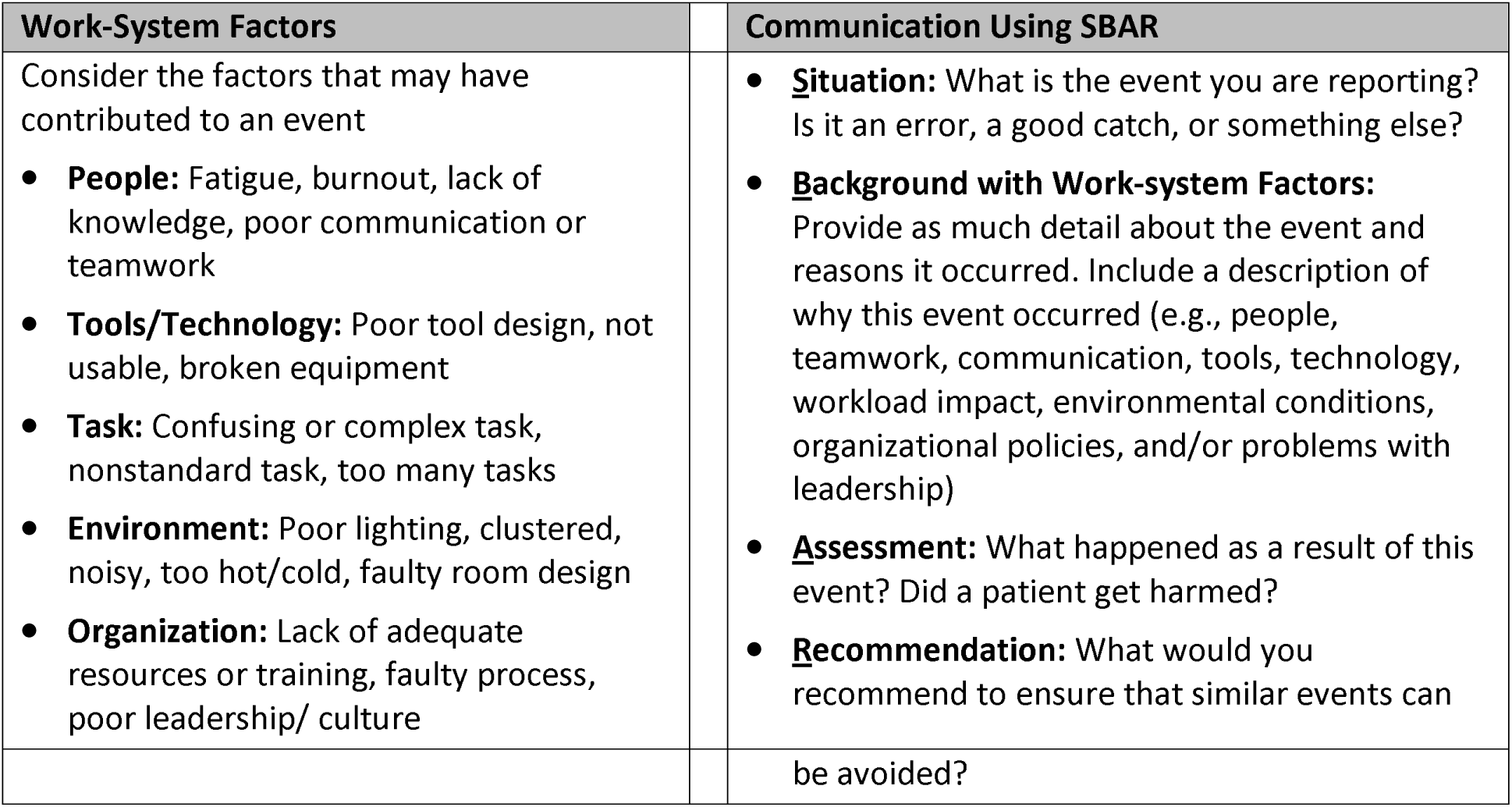
SAFE Loop Training in Incident Reporting: Key Messages.

| Work-System Factors | Communication Using SBAR |
| --- | --- |
| Consider the factors that may have contributed to an event<br>• **People:** Fatigue, burnout, lack of knowledge, poor communication or teamwork<br>• **Tools/Technology:** Poor tool design, not usable, broken equipment<br>• **Task:** Confusing or complex task, nonstandard task, too many tasks<br>• **Environment:** Poor lighting, clustered, noisy, too hot/cold, faulty room design<br>• **Organization:** Lack of adequate resources or training, faulty process, poor leadership/ culture | • **Situation:** What is the event you are reporting? Is it an error, a good catch, or something else?<br>• **Background with Work-system Factors:** Provide as much detail about the event and reasons it occurred. Include a description of why this event occurred (e.g., people, teamwork, communication, tools, technology, workload impact, environmental conditions, organizational policies, and/or problems with leadership)<br>• **Assessment:** What happened as a result of this event? Did a patient get harmed?<br>• **Recommendation:** What would you recommend to ensure that similar events can |

|  |  |  |
| --- | --- | --- |
|  |  | be avoided? |

To deploy the training program on the pilot unit, we loaded it into an online platform, obtained a list of nursing staff, and enrolled the nurses in the mandatory training. Nursing unit leaders monitored the nurses’ completion and provided encouragement where necessary.

#### Prompting Incident Reporting

During the pilot testing period, we periodically revisited the nursing huddles to remind nurses to report the Target Events, monitored the medication safety incidents being reported, and let nurses know that we received and appreciated the reports they submitted. We also checked in often with the nursing unit leaders about what nurses were saying about the training, Target Event, and other aspects of the SAFE Loop.

#### Investigative Procedures

We designed Target Event follow-up procedures to be brief but thorough. First, the champion reviewed the medication-related incident reports submitted during the six months preceding and during the pilot test. They considered whether each incident might be relevant to the Target Event, identified contributing work-system factors in the narrative, and used the Human Factors Analysis and Classification System for Healthcare (HFACS-Healthcare) to categorize them [73].

Concurrently, the champion and two other research team members interviewed nursing staff on the unit over two 90-minute sessions. During each session, nursing unit leaders directed one to three nurses at a time to a conference room on the unit, where we explored their thoughts about the chosen Target Event and contributing factors. Each nurse participated in the interviews for about 10-15 minutes, without being away from patient care or disrupting work on the unit for too long. As information about the Target Event and contributing work-system factors came to light, the champion and research team members identified additional types of clinicians with the potential to observe or contribute to the Target Event (e.g., pharmacists or physicians), and administrative leaders who oversaw departments that contributed to the work-system environment for unit nurses. We then conducted brief interviews with these individuals about the Target Events, starting with open-ended questions and gradually seeking feedback on what we had heard about contributing work-system factors.

At the same time, we obtained information about the history of the Target Event at the institution from the Pharmacist Manager for Medication Safety and the Chief Patient Safety Officer on our research team. They shared the results of prior investigative activities, shed light on contributing work-system factors, and provided insight into potential strategies to mitigate them and barriers.

Since a medication safety event at our hospital was likely to occur at other institutions, too, we considered external information, to leverage any previous lessons learned. The champion conducted a brief, directed (non-systematic) literature review to identify prior publications. The champion contacted the study’s partners at two AHRQ-designated Patient Safety Organizations (PSOs) to see what prior investigative activities and organizational activities or reports might have been relevant to the Target Event. The PSOs included the Hospital Quality Institute (engaged with patient safety and incident reporting at hospitals in California), and the Institute for Safe Medication Practices (engaged with medication safety and incident reporting at hospitals nationally).

To conclude the follow-up activities, the champion created an Ishikawa Diagram for the Target Event, listing the specific contributing factors identified via incident reports and interviews with nurses or other hospital stakeholders.

#### Producing a Mitigation Plan

Next, we met with the nursing unit leaders, the Pharmacist Manager for Medication Safety, the Chief Patient Safety Officer, hospital nursing leaders, and leaders of other relevant departments as needed. We discussed the Target Event and the contributing factors, including those that were reported most often, appeared most preventable, and/or posed the most significant risk. We also discussed relevant institutional history and any information from external sources.

Nursing leaders ultimately decided what work-system factors to prioritize for mitigation and implemented the mitigation plan, based on their understanding of the issues and what changes may be feasible. Once the strategy was selected, the research team drafted a two-page mitigation plan, reiterating the Target Event, describing the contributing factors (including the Ishikawa Diagram), steps we had taken in the work, any outside information we identified, and the nursing unit’s final mitigation plan. After the nursing unit director approved the mitigation plan, we emailed it to all nurses on the unit.

### Step (C) Structured Pilot Testing

As a pilot study, the focus was on maximizing feasibility and acceptability to nursing unit leaders and nursing staff, and on refining the intervention to enhance effectiveness. Analyses were not powered to test for statistical significance.

#### Incident Reporting Practices

We measured the number of weekly incident reports submitted on the units during six months of the pilot test (June 2, 2022, to December 12, 2022) and a prior six-month period (December 1, 2021, to June 1, 2022). Dual reviewers examined and reached a consensus on whether each report included work-system factors in the narrative and whether it pertained to the unit’s selected Target Event; the team adjudicated any ties. We also used severity scores assigned by the reporter to identify near misses (NCC MERP categories A to C) [65, 66, 74].

#### Surveys of Nurses

Before and after implementation, we surveyed the front-line nurses by email, with 10 items described below (**Appendix**). To promote the survey, we visited the nursing units with snacks and provided encouragement. Additionally, nurses who completed each survey were entered into a lottery for Apple AirPods.

From the AHRQ Hospital Survey on Patient Safety (SOPS) Culture™, Version 2.0 (English) [75], we derived two composite measures, Communication about Error (3 items) and Reporting Patient Safety Events (2 items), and two individual items, Blame-free Culture and Incident Report Submissions. We made minor changes in item wording to enhance specificity (e.g., changing “we” to “nurses” and “event reports” to “incident reports”). We also created a novel composite measure, Confidence in Reporting (2 items), and a novel individual measure, Time per Report.

Most individual items were scored on a scale from 1 (strongly disagree or similar) to 5 (strongly agree or similar). Composite scores are calculated as the percent of positive responses (agree or strongly agree), ranging from 0 to 100% (higher is better). Incident Report Submissions included five categories (0 to 4+ reports). Time per Report included 5-minute increments up to 90 minutes.

#### Interviews with Nurses

After implementation of the SAFE Loop, we conducted one-on-one in-person interviews with three nursing unit leaders and three frontline nurses. We used a semi-structured interview guide with open-ended questions and follow-up probes, guided by the Consolidated Framework for Implementation Research and key publications on information to report about quality improvement interventions [76–79]. Questions were designed to elicit respondents’ views regarding contextual factors, implementation details, and other information recommended for the reporting of interventions related to quality and safety. Pairs of researchers trained in qualitative analysis analyzed interview transcripts in Dedoose using a combination of content analysis and qualitative inquiry, allowing them to identify key themes related to the nurses’ experiences and perceptions.

## Results

### Step (A) Proof-of-concept Testing

#### Feasibility of Implementation

Nursing unit leaders and front-line nurses were willing and able to select a Target Event. The nurses submitted several incident reports relevant to the Target Events. The unit leaders and nursing staff were receptive to discussing findings from the incident reports and mini-root-cause analyses. Finally, it was feasible to work with the nursing units to implement tests of change and disseminate mitigation plans.

#### Reporting Practices

Before the pilot test on these two nursing units, an average of four incident reports were submitted per week and 6.25% represented near misses. During the pilot test, an average of 10 reports were submitted per week and 47% of reports addressed near misses.

#### Surveys of Nurses

Before implementation, the Culture of Safety composite measure was 91% on one unit and 75% on the other. These percentages were 94% and 79%, respectively, after implementation.

#### Interviews with Nurses

Feedback from unit leaders was strongly favorable: “SAFE Loop was an effective test of change as it engaged front-line staff to voice input and concerns to prevent near misses. It improved their workflow processes in terms of helping eliminate workarounds or delays. Staff had an increased feeling of empowerment.”

One front-line nurse commented, “We saw the value of these rounds. It made our job easier and more efficient, and we could see the long-term efforts of putting in the near misses now.” Another said, “[The advantages include] more reporting of ‘near misses’ and ensuring all staff receive follow-up reports.”

### Step (B) Refined Intervention Design and Implementation

#### Obtaining Input from Nurses and Selecting a Target Event

During SAFE Loop implementation in the structured pilot study, unit directors initially proposed the Target Event as “challenges using the Pyxis safely.” Discussions with nursing unit directors and nurses revealed that this encompassed a wide variety of concerns. For the SAFE Loop, the Target Event needed to occur fairly often (e.g., at least once per month), be preventable, and be clearly understood by front-line nurses. Consequently, the study team and nursing unit leaders narrowed down a list of more specific issues related to automated dispensing machines and reframed the Target Event as “wrong medication or wrong patient errors involving the Pyxis.” Further clarifications of the event definition were necessary during the investigation and mitigation planning activities to produce a focused and feasible set of actions in the nursing unit.

#### Training in Enhanced Reporting

During the cognitive testing of the draft training program, nurses reported that they value information on how to submit more informative incident reports, since they receive limited training as part of nursing education. Testers found the training to be easily understandable and useful, and reported that they could complete the training in 15-20 minutes.

At the beginning of implementation on the pilot unit, 20 of 26 full-time nurses (77%) completed the online training program.

#### Prompting Incident Reporting

During the investigative interviews with nurses (below), we learned that nurses might collectively recognize a high-priority safety issue on the unit but not spontaneously submit incident reports about it. In this case, the Target Event—the interruptions while using the automated dispensing machine—occurred so often as to seem a routine part of care rather than a reportable incident. As such, the interviews reinforced the submission of reports on the Target Events.

#### Investigative Procedures

During the implementation process, we learned that nurses were generally eager to share their thoughts about the Target Event during the brief interviews. While speaking to individual nurses was helpful, nurses became more engaged when we talked with to two to three at a time, and discussions among the nurses appeared to stimulate more and richer insights. We also learned that identifying and understanding contributing factors were much easier to accomplish via interviews, rather than from incident reports.

The investigation yielded four incident reports directly relevant to the Target Event, 10 interviews (six with nurses on the unit, and four with other informants), 12 relevant peer-reviewed publications, and eight resources shared by the Patient Safety Organizations.

The incident reports and interviews yielded 21 unique contributing factors for this Target Event (Figure 2, Ishikawa Diagram). Of these 21 factors, 17 were derived from nursing interviews alone, one from an incident report alone, and three from both nursing interviews and incident reports. The contributing factor discussed most often was distractions or interruptions while using the Pyxis machine, which was located in an open area by the nurse station.

**Figure 2:**
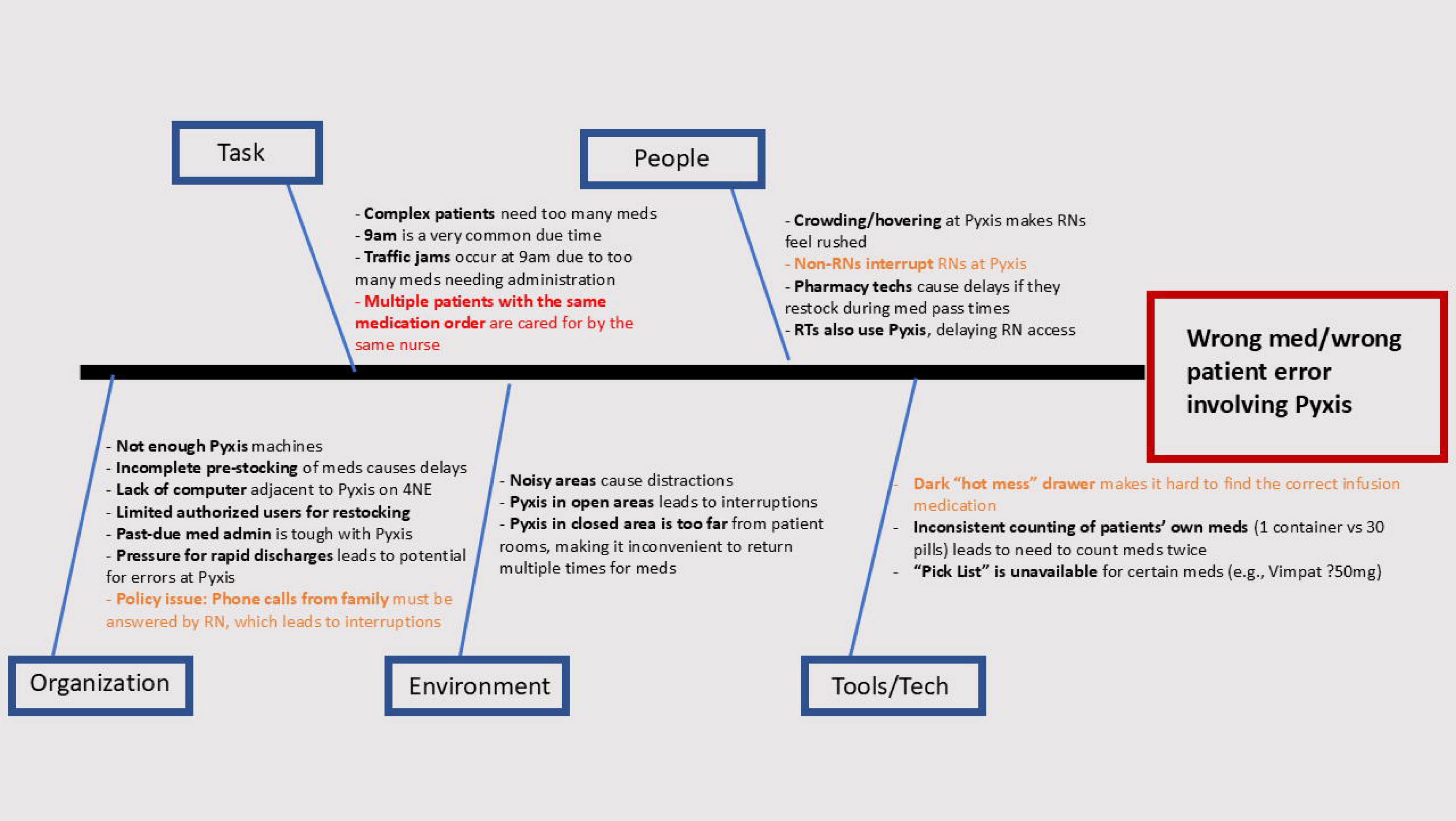
Results of Structured Pilot Test of SAFE Loop Intervention: Contributing Work-system Factors Identified by Investigation (Ishikawa Diagram) Legend: Black text signifies contributing factors identified only during interviews, Red text signifies contributing factors identified only from incident reports, and orange text signifies contributing factors identified and/or characterized by both data sources.]

#### Producing a Mitigation Plan

The selection of the Target Event and subsequent investigation illustrate that nurses recognized numerous threats to patient safety on the unit and that a threat can have many diverse contributing work-system factors. As such, nursing leaders in the unit had to make pragmatic decisions about prioritizing these factors. They chose to focus on distractions while using the automated dispensing machine.

To address this, the nursing directors selected two mitigation strategies: (1) a new “quiet zone” sign on the automated dispensing machine to alert people not to interrupt nurses while they are retrieving medications, and (2) a new policy to defer all non-urgent calls (i.e., from patients’ families) during morning medication administration periods to reduce interruptions while using the automated dispensing machine. The nursing unit directors immediately implemented these changes. Step (C) Structured Pilot Testing:

#### Reporting Practices

In the six months before the pilot test, nurses submitted 48 medication-related incident reports, including 18 (37.5%) related to the Pyxis. Of 28 events for which severity was reported, 22 (79%) were near misses (7 unsafe conditions, three did not reach the patient, 12 did not harm). Across the 48 reports, nurses described 53 contributing work-system factors (Table 4).

**Table 4:** Results of Structured Pilot Test of SAFE Loop Intervention: Contributing Work-system Factors Obtained from Incident Report Narratives.

| HFACS-Healthcare Tier | Contributing Factor Category | Baseline Reports, N (%) | Pilot Test Reports, N (%) |
| --- | --- | --- | --- |
| Organizational Influences | Organizational Climate | 0 | 0 |
|  | Operational Process | 0 | 0 |
|  | Resource Management | 0 | 0 |
|  | *Subtotals* | 0 | 0 |
| Supervisory Factors | Inadequate Supervision | 0 | 1 |
|  | Planned Inappropriate Operations | 0 | 0 |
|  | Failed to Correct Problem | 0 | 1 |
|  | Supervisory Violations | 0 | 0 |
|  | *Subtotals* | 0 | 2 |
| Preconditions for Unsafe Acts | Physical Environment | 0 | 0 |
|  | Tools/Technology | 2 | 0 |
|  | Task | 1 | 3 |
|  | Mental State | 1 | 0 |
|  | Physiological State | 0 | 0 |
|  | Fitness for Duty | 0 | 0 |
|  | Communication | 7 | 3 |
|  | Coordination | 3 | 3 |
|  | Leadership | 0 | 0 |
|  | *Subtotals* | 14 | 9 |
| Unsafe Acts | Decision errors | 6 | 7 |
|  | Skill-Based Errors | 26 | 17 |
|  | Perceptual Errors | 0 | 0 |
|  | Routine Violations | 0 | 3 |
|  | Exceptional Violations | 1 | 0 |
|  | *Subtotals* | 33 | 27 |
|  | **Total Incident Reports** | 48 | 41 |
|  | **Total Contributing Factors** | 53 | 39 |

During the 6-month pilot test, nurses submitted 41 medication-related incidents, including 8 (19.5%) related to the Pyxis. Of 30 events for which severity was reported, 25 (83%) were near misses (4 unsafe conditions, four did not reach the patient, 17 no harm). Across the 41 reports, nurses described 39 contributing work-system factors.

#### Surveys of Nurses

Survey responses were generally similar before and after SAFE Loop implementation (**Table 5**). The Time per Report rose from 10 minutes to 15.5 minutes.

**Table 5:** Results of Investigation Structured Pilot Test on One Nursing Unit: Attitudes of Front-line Nurses Toward Reporting.

|  | **Baseline Survey** | **Survey after Pilot Test** |
| --- | --- | --- |
| **Response Rate** | 22 of 54 (40.7%) | 29 of 59 (49.2%) |
| **Composite Measures** |  |  |
| Confidence in Reporting, % Positive Responses | 70.5% | 82.7% |
| Communication About Error, % Positive Responses | 74.2% | 70.1% |
| Reporting Patient Safety Events, % Positive Responses | 70.5% | 69.0% |
| **Individual Items** |  |  |
| Blame-free Culture, % Positive Responses | 86.4% | 82.8% |
| Incident Report Submissions, 0-4+ |  |  |
| None | 15 (68.2%) | 19 (65.5%) |
| 1 | 5 (22.7%) | 5 (17.2%) |
| 2 | 1 (4.5%) | 2 (6.9%) |
| 3 | 1 (4.5%) | 1 (3.4%) |
| 4 or more | 0 (0%) | 2 (6.9%) |
| Time per Report, Mean Minutes (SD)* | 10 (5.8) | 15.5 (10.0) |

\*Excludes missing responses

#### Interviews with Nurses

Nurses’ responses revealed that, before SAFE Loop implementation, attitudes towards incident reporting were generally favorable. However, nurses were uncertain about what changes were made to improve safety in response to incident reporting.

After SAFE Loop implementation, nurses voiced positive opinions of the potential for an increase in incident reporting to improve medication safety, and a nursing leader reported a high level of receptivity and investment from nurses in reporting the Target Event. See Box 1.

##### Box 1

###### Comments Related to Incident Reporting on the Nursing Unit <u>before</u> SAFE Loop Implementation

- All nurses reported it was their responsibility to submit incident reports because:

- *“[Nurses are] the first in line to see [a] reportable event.”*
- The culture of safety was generally positive:

- *“If we see something then we say something. We can tell the charge nurse or they really encourage us to just report anything, whether it’s medication related or [something else].”*
- However, nurses’ opinions were mixed about whether they received follow-up information after submitting a report.

- *“I mean it really depends. So far, every time we’ve submitted forms, I’ve gotten the email [confirming that the incident report was received by the reporting system] for sure. And then just depending on the situation, I’ll have management come and say, ‘Hey, just so you know, we got the form and we’re looking into it’ kind of thing.”*
- *“Really, once the file has been reported, we don’t really hear about what happens.”*
- Nurses were less certain about whether submitting an incident report leads to changes in medication safety on the unit:

- *“I’d like to think that leadership sees it and does things about it. And sometimes I feel like they do, sometimes they talk about what had happened in our huddle, but it’s not for everything. So, I really don’t know how effective the incident reporting is.”*
- *“I’m not sure because I don’t know if people, or who is reporting medication errors and what gets done about them. So maybe it’s effective, but I don’t see how it’s affected me before.”*
- *“And have you found that that med report incident reporting is effective at improving medication safety? Yes. For pharmaceuticals.”*

###### Comments Related to Incident Reporting on the Nursing Unit after SAFE Loop Implementation

- Nurses voiced a potential for an increase in incident reporting to improve medication safety, particularly the combination of reporting via the electronic interface and in-person to charge nurses.
- *“[I think] people learn from their mistakes, people learn through mistakes… So, if the errors are recorded properly, then I believe they’ll learn about it. And then, we can make plans on how we can further prevent the same incident from happening, and then also study how this kind of incidents happen, if they have this multiple times.”*
- One nursing leader reported a high level of receptivity and investment from nurses in reporting the Target Event:
- *“I think they have it down. They have it down. Okay. Yeah, they’re very invested and making sure that when the whole system started, the whole CS-Safe entry process started, they saw it as a way of maybe potentially falling on someone’s desk who was removed from the clinical aspects. We may do nothing with it. So, we weren’t seeing the real meaty, detailed CS-Safe events, but now we’re seeing more and a lot more of the entries being made and capturing. We try to tell them, remove the emotion, just get to the facts. And we’re seeing a lot more of that. So, it’s slowly getting less personal and getting to the core of what the issue was. So, we’re seeing they are more receptive.”*

## Discussion

This analysis involved preliminary testing and iterative refinement of the Safety Action Feedback and Engagement (SAFE) Loop, a novel multifaceted intervention designed to overcome several limitations of hospital incident reporting systems and transform them into more valuable tools for improving patient safety. We quickly and easily implemented the intervention on two nursing units in a small initial proof-of-concept test, suggesting good feasibility. Nurses voiced substantial enthusiasm for the SAFE Loop approach, suggesting good acceptability. After specifying the SAFE Loop intervention and implementation activities in detail, incorporating ongoing feedback from nurses, and making many refinements, we conducted a structured pilot test of the full intervention using comprehensive mixed evaluation methods. The structured pilot test reaffirmed feasibility, including for specific activities like selecting a Target Event together with nursing units, training nurses in the SAFE Loop enhanced reporting methods, engaging nurses via visits to twice-daily “huddles” at shift change, conducting investigative interviews with front-line nurses about the selected Target Event, integrating information about the Target Event from diverse sources, developing a mitigation plan with the nursing unit leaders, and closing the feedback loop by distributing the mitigation plan to nurses on the unit. The analysis also demonstrated the feasibility of our evaluation methods, including evaluating the information incident reports capture about contributing work-system factors, and surveying nurses’ attitudes toward reporting. Collaborative interactions with nursing unit leaders and front-line nurses throughout the project reaffirmed acceptability, as did qualitative interviews performed after the pilot. As a whole, the testing and refinements laid the groundwork for initial efficacy testing via a randomized controlled trial.

The present analysis did not involve statistical testing because sample sizes lacked sufficient power and underpowered pilot tests to assess efficacy have high rates of false positives and false negatives relative to fully powered trials [80]. Our results, nonetheless, offer a few initial surprises. First, during the Target Event selection process, we learned that nurses might collectively recognize a high-priority safety issue on the unit, but may not spontaneously submit many incident reports about it. In this case, the Target Event—the interruptions during use of the automated dispensing machine— occurred so often as to seem a routine part of care, rather than a reportable incident.

Second, of 21 different work-system factors that nurses identified as contributing to the selected Target Event, we detected only one from incident reporting alone. In contrast, brief investigative interviews with 10 nurses discovered 20 more work-system factors. Three of the 20 factors discovered via interviews were also mentioned in incident reports, but we obtained much richer information during interviews due to the ability to ask follow-up questions, invite detailed descriptions, and explore nuances and diverse opinions. We suspect that many nurses felt more comfortable disclosing sensitive information during unrecorded face-to-face discussions with neutral third parties, than they did about putting this information in a written report that their supervisors would read.

A third unanticipated finding was that, while nurses chose a problem involving the Pyxis machine as their Target Event, they submitted less than half as many Pyxis incidents during the structured pilot test than in the preceding six months. Our team and the unit champion appeared frequently on the nursing unit to prompt nurses to report instances of the Target Event, but this may have backfired by offering an alternative way of sharing insights, i.e., via the interviews.

We will examine these issues in greater depth in our pragmatic randomized controlled trial testing the efficacy of the SAFE Loop intervention (1R01HS027455, clinical trial registration number) [51, 81]. To our knowledge, no prior randomized controlled trials have examined interventions designed to improve hospital incident reporting systems. Moreover, the SAFE Loop is unique in that it is designed to simultaneously overcome multiple limitations of these reporting systems. Most prior reporting interventions have focused on a single detail, such as educating nurses and physicians about reporting, or changing from paper to electronic incident reports [81]. The SAFE Loop’s emphasis on clinician engagement is crucial because incident reporting is voluntary. After all, safety emerges from the summary of countless small actions by clinicians, and nurses can provide rich insights into threats to patient safety. Critically, the SAFE Loop emphasizes the fundamental purpose of incident reports as qualitative tools for gaining insight into the causes of critical events, based on much more successful applications of incident reporting in other high-risk industries. We are unaware of prior efforts to train nurses to recognize and report work-system factors. Also, the SAFE Loop proposes standardizing procedures for leveraging the information in incident reports, closing what experts call the “safety action feedback loop” [34].

Despite these advantages, our present work has several potential limitations. We tested the SAFE Loop at one large urban academic center with a strong commitment to safety, so adaptation to and evaluation in other contexts will be needed. Implementation of the SAFE Loop will likely differ across nursing units and over time because the intervention involves tailoring to local needs, and the engagement of nursing unit leaders and nurses will undoubtedly vary. The types of safety problems that SAFE Loop could solve will be influenced by its current focus on reporting by nurses. Front-line nurses may have a skewed perspective relative to an objective process for prioritizing patient safety events (e.g., based on frequency, severity, and preventability) or patient preferences. Nonetheless, many safety problems are inherently local, such as poor communication and coordination of care, and nursing units would benefit from new strategies for solving their local problems. The intervention could be extended to physicians and other clinicians in the future. The SAFE Loop is resource intensive, requiring the efforts of nursing unit leaders, a safety champion, front-line nurses, and many others. As such, it may be best used to solve only a few safety problems in a given setting per year. As with many multifaceted interventions designed to improve the quality of care, it will be hard to isolate which components of the SAFE Loop are most important to any future successes.

In conclusion, this work demonstrated the feasibility and acceptability of a novel intervention, the SAFE Loop, for hospital nurses. An ongoing trial is testing its effects on reporting practices, nurses’ attitudes toward reporting, medication event rates and exploring nurses’ perspectives on implementation.

## Supporting information

Appendix survey

IRB approval letter

## Data Availability

Data cannot be shared publicly because they contain patient and clinician identifiers and represent sensitive information for hospitals.

## Notes

**Funding Disclosure**: The Agency for Healthcare Research and Quality (AHRQ) funded this work (R01HS027455). Study investigators have copyrighted certain materials resulting from this work. Dr. Nuckols also receives funding from NIH/NCATS (UL1TR001881) and Dr. Berdahl is supported by an AHRQ K08 Career Development Award (K08HS029534).

### Competing Interest Statement

The authors have declared no competing interest.

### Clinical Trial

ClinicalTrials.gov Number NCT05381441

### Clinical Protocols

https://doi.org/10.1016/j.conctc.2023.101192

### Funding Statement

The Agency for Healthcare Research and Quality (AHRQ) funded this work (R01HS027455). Study investigators have copyrighted certain materials resulting from this work. Dr. Nuckols also receives funding from NIH/NCATS (UL1TR001881) and Dr. Berdahl is supported by an AHRQ K08 Career Development Award (K08HS029534).

### Author Declarations

Cedars-Sinai Institutional Review Board, Office of Research Compliance & Quality Improvement

## References

1. Eldridge N, Wang Y, Metersky M, Eckenrode S, Mathew J, Sonnenfeld N, et al. Trends in Adverse Event Rates in Hospitalized Patients, 2010-2019. Jama. 2022;328(2):173–83. doi: 10.1001/jama.2022.9600. PubMed PMID: 35819424; PubMed Central PMCID: PMCPMC9277501.

2. Baines R, Langelaan M, De Bruijne M, Spreeuwenberg P, Wagner C. How effective are patient safety initiatives? A retrospective patient record review study of changes to patient safety over time. BMJ Quality and Safety. 2015. doi: 10.1136/bmjqs-2014-003702.

3. National and State Healthcare-Associated Infections Progress Report: Centers for Disease Control and Prevention, Atlanta, GA; 2017. Available from: https://www.cdc.gov/hai/data/portal/progress-report.html.

4. Bates DW, Levine DM, Salmasian H, Syrowatka A, Shahian DM, Lipsitz S, et al. The Safety of Inpatient Health Care. N Engl J Med. 2023;388(2):142–53. doi: 10.1056/NEJMsa2206117. PubMed PMID: 36630622.

5. Landrigan CP, Parry GJ, Bones CB, Hackbarth AD, Goldmann DA, Sharek PJ, et al. Temporal Trends in Rates of Patient Harm Resulting from Medical Care. N Engl J Med. 2010. doi: 10.1056/NEJMsa1004404.

6. Sunshine JE, Meo N, Kassebaum NJ, Collison ML, Mokdad AH, Naghavi M. Association of Adverse Effects of Medical Treatment With Mortality in the United States: A Secondary Analysis of the Global Burden of Diseases, Injuries, and Risk Factors Study. JAMA network open. 2019;2(1):e187041-e. doi: 10.1001/jamanetworkopen.2018.7041.

7. Panagioti M, Khan K, Keers RN, Abuzour A, Phipps D, Kontopantelis E, et al. Prevalence, severity, and nature of preventable patient harm across medical care settings: systematic review and meta-analysis. Bmj. 2019;366:l4185. Epub 20190717. doi: 10.1136/bmj.l4185. PubMed PMID: 31315828; PubMed Central PMCID: PMCPMC6939648.

8. de Vries EN, Ramrattan MA, Smorenburg SM, Gouma DJ, Boermeester MA. The incidence and nature of in-hospital adverse events: a systematic review. Qual Saf Health Care. 2008;17(3):216–23. doi: 10.1136/qshc.2007.023622. PubMed PMID: 18519629; PubMed Central PMCID: PMCPMC2569153.

9. Jehring J, Heinrich HW. Industrial Accident Prevention: A Scientific Approach. Industrial and Labor Relations Review. 2006. doi: 10.2307/2518508.

10. Gordon F RD. The cost to Britain of workplace accidents and work-related ill health in 1995/96 Sudbury, Suffolk, UK: Health and Safety Executive; 1999 [cited 2019 05/31]. Available from: http://www.hse.gov.uk/pUbns/priced/hsg101.pdf.

11. Johnson CW. A Handbook of Incident and Accident Reporting. Glasgow: Glasgow University Press; 2003.

12. Flanagan JC. The critical incident technique. Psychol Bull. 1954;51(4):327–58. doi: 10.1037/h0061470. PubMed PMID: 13177800.

13. Reporting Patient Safety Events: Agency for Healthcare Research and Quality, Rockville, MD; 2019. Available from: https://psnet.ahrq.gov/primers/primer/13/reporting-patient-safety-events.

14. Woods D. Improving the reporting and analysis of incidents. Enhancing Patient Safety and Reducing Errors in Health Care, Chicago. 1999.

15. Vincent C. Clinical risk management: Enhancing patient safety: BMJ Publishing group; 2001.

16. Nuckols TK, Bell DS, Liu H, Paddock SM, Hilborne LH. Rates and types of events reported to established incident reporting systems in two US hospitals. Qual Saf Health Care. 2007;16(3):164–8. Epub 2007/06/05. doi: 10.1136/qshc.2006.019901. PubMed PMID: 17545340; PubMed Central PMCID: PMCPMC2464990.

17. Gampetro PJ, Nickum A, Schultz CM. Perceptions of U.S. and U.K. Incident Reporting Systems: A Scoping Review. J Patient Saf. 2024;20(5):360–9. Epub 20240429. doi: 10.1097/pts.0000000000001231. PubMed PMID: 38682884.

18. Burlison JD, Quillivan RR, Kath LM, Zhou Y, Courtney SC, Cheng C, Hoffman JM. A Multilevel Analysis of U.S. Hospital Patient Safety Culture Relationships With Perceptions of Voluntary Event Reporting. J Patient Saf. 2020;16(3):187–93. doi: 10.1097/pts.0000000000000336. PubMed PMID: 27820722; PubMed Central PMCID: PMCPMC5415419.

19. Hamed MMM, Konstantinidis S. Barriers to Incident Reporting among Nurses: A Qualitative Systematic Review. West J Nurs Res. 2022;44(5):506–23. Epub 20210317. doi: 10.1177/0193945921999449. PubMed PMID: 33729051.

20. Mitchell I, Schuster A, Smith K, Pronovost P, Wu A. Patient safety incident reporting: A qualitative study of thoughts and perceptions of experts 15 years after ‘To Err is Human’. BMJ Quality and Safety. 2016. doi: 10.1136/bmjqs-2015-004405.

21. Evans SM, Berry JG, Smith BJ, Esterman A, Selim P, O’Shaughnessy J, DeWit M. Attitudes and barriers to incident reporting: A collaborative hospital study. Quality and Safety in Health Care. 2006. doi: 10.1136/qshc.2004.012559.

22. Archer S, Hull L, Soukup T, Mayer E, Athanasiou T, Sevdalis N, Darzi A. Development of a theoretical framework of factors affecting patient safety incident reporting: A theoretical review of the literature. 2017.

23. Feeser VR, Jackson AK, Savage NM, Layng TA, Senn RK, Dhindsa HS, et al. When Safety Event Reporting Is Seen as Punitive: “I’ve Been PSN-ed!”. Ann Emerg Med. 2021;77(4):449–58. Epub 20200815. doi: 10.1016/j.annemergmed.2020.06.048. PubMed PMID: 32807540.

24. Cullen DJ, Bates DW, Small SD, Cooper JB, Nemeskal AR, Leape LL. The incident reporting system does not detect adverse drug events: a problem for quality improvement. The Joint Commission journal on quality improvement. 1995. doi: 10.1016/S1070-3241(16)30180-8.

25. Sari ABA, Sheldon TA, Cracknell A, Turnbull A. Sensitivity of routine system for reporting patient safety incidents in an NHS hospital: Retrospective patient case note review. 2007.

26. Hibbert PD, Molloy CJ, Schultz TJ, Carson-Stevens A, Braithwaite J. Comparing rates of adverse events detected in incident reporting and the Global Trigger Tool: a systematic review. Int J Qual Health Care. 2023;35(3). doi: 10.1093/intqhc/mzad056. PubMed PMID: 37440353; PubMed Central PMCID: PMCPMC10367579.

27. Cohen TN, Francis SE, Wiegmann DA, Shappell SA, Gewertz BL. Using HFACS-Healthcare to Identify Systemic Vulnerabilities During Surgery. American Journal of Medical Quality. 2018:1062860618764316.

28. Nuckols TK, Bell DS, Paddock SM, Hilborne LH. Comparing process- and outcome-oriented approaches to voluntary incident reporting in two hospitals. Jt Comm J Qual Patient Saf. 2009;35(3):139–45. Epub 2009/03/31. PubMed PMID: 19326805.

29. Nuckols TK, Bell DS, Paddock SM, Hilborne LH. Contributing factors identified by hospital incident report narratives. BMJ Quality & Safety. 2008;17(5):368–72.

30. Scott J, Dawson P, Heavey E, De Brún A, Buttery A, Waring J, Flynn D. Content Analysis of Patient Safety Incident Reports for Older Adult Patient Transfers, Handovers, and Discharges: Do They Serve Organizations, Staff, or Patients? J Patient Saf. 2021;17(8):e1744–e58. doi: 10.1097/pts.0000000000000654. PubMed PMID: 31790011; PubMed Central PMCID: PMCPMC8612895.

31. Battles JB, Stevens DP. Adverse event reporting systems and safer healthcare. Quality and Safety in Health Care. 2009.

32. Nuckols TK. Incident Reporting: More Attention to the Safety Action Feedback Loop, Please: Agency for Healthcare Research and Quality; 2011. Available from: https://psnet.ahrq.gov/perspective/incident-reporting-more-attention-safety-action-feedback-loop-please.

33. Macrae C. The problem with incident reporting. BMJ Qual Saf. 2016;25(2):71–5. Epub 2015/09/09. doi: 10.1136/bmjqs-2015-004732. PubMed PMID: 26347519.

34. Benn J, Koutantji M, Wallace L, Spurgeon P, Rejman M, Healey A, Vincent C. Feedback from incident reporting: information and action to improve patient safety. Qual Saf Health Care. 2009;18(1):11–21. Epub 2009/02/11. doi: 10.1136/qshc.2007.024166. PubMed PMID: 19204126.

35. Jäger C, Mohr G, Gökcimen K, Navarini A, Schwendimann R, Müller S. Critical incident reporting over time: a retrospective, descriptive analysis of 5493 cases. Swiss Med Wkly. 2021;151:w30098. Epub 20211228. doi: 10.4414/smw.2021.w30098. PubMed PMID: 34964949.

36. McNiven B, Wu T, Brown AD. Novel Telephone-Based Interactive Voice Response System for Incident Reporting. Jt Comm J Qual Patient Saf. 2021;47(12):809–13. Epub 20210925. doi: 10.1016/j.jcjq.2021.09.010. PubMed PMID: 34732307.

37. Kapur N, Parand A, Soukup T, Reader T, Sevdalis N. Aviation and healthcare: a comparative review with implications for patient safety. JRSM Open. 2016. doi: 10.1177/2054270415616548.

38. Erickson SM, Wolcott J, Corrigan JM, Aspden P. Patient safety: achieving a new standard for care: National Academies Press; 2003.

39. Skutezky T, Small SS, Peddie D, Balka E, Hohl CM. Beliefs and perceptions of patient safety event reporting in a Canadian Emergency Department: a qualitative study. Cjem. 2022;24(8):867–75. Epub 20221107. doi: 10.1007/s43678-022-00400-2. PubMed PMID: 36344901; PubMed Central PMCID: PMCPMC9763130.

40. Goekcimen K, Schwendimann R, Pfeiffer Y, Mohr G, Jaeger C, Mueller S. Addressing Patient Safety Hazards Using Critical Incident Reporting in Hospitals: A Systematic Review. J Patient Saf. 2023;19(1):e1–e8. Epub 20220820. doi: 10.1097/pts.0000000000001072. PubMed PMID:35985209; PubMed Central PMCID: PMCPMC9788933.

41. Oyibo K, Gonzalez PA, Ejaz S, Naheyan T, Beaton C, O’Donnell D, Barker JR. Exploring the Use of Persuasive System Design Principles to Enhance Medication Incident Reporting and Learning Systems: Scoping Reviews and Persuasive Design Assessment. JMIR Hum Factors. 2024;11:e41557. Epub 20240321. doi: 10.2196/41557. PubMed PMID: 38512325; PubMed Central PMCID: PMCPMC10995789.

42. Leape LL, Brennan TA, Laird N, Lawthers AG, Localio AR, Barnes BA, et al. The nature of adverse events in hospitalized patients. Results of the Harvard Medical Practice Study II. N Engl J Med. 1991;324(6):377–84. Epub 1991/02/07. doi: 10.1056/nejm199102073240605. PubMed PMID:1824793.

43. Thomas EJ, Studdert DM, Burstin HR, Orav EJ, Zeena T, Williams EJ, et al. Incidence and types of adverse events and negligent care in Utah and Colorado. Med Care. 2000;38(3):261–71. Epub 2000/03/16. PubMed PMID: 10718351.

44. Patient Safety Network: Medication Errors and Adverse Drug Events: Agency for Healthcare Research and Quality, Rockville, MD; 2019. Available from: https://psnet.ahrq.gov/primers/primer/23/Medication-Errors-and-Adverse-Drug-Events.

45. Hauck K, Zhao X. How dangerous is a day in hospital? A model of adverse events and length of stay for medical inpatients. Med Care. 2011;49(12):1068–75. Epub 2011/09/29. doi: 10.1097/MLR.0b013e31822efb09. PubMed PMID: 21945976.

46. Bates DW, Cullen DJ, Laird N, Petersen LA, Small SD, Servi D, et al. Incidence of adverse drug events and potential adverse drug events. Implications for prevention. ADE Prevention Study Group. JAMA. 1995;274(1):29–34. Epub 1995/07/05. PubMed PMID: 7791255.

47. Hawkins J, Madden K, Fletcher A, Midgley L, Grant A, Cox G, et al. Development of a framework for the co-production and prototyping of public health interventions. BMC Public Health. 2017;17(1):689. Epub 20170904. doi: 10.1186/s12889-017-4695-8. PubMed PMID: 28870192;PubMed Central PMCID: PMCPMC5583990.

48. Czajkowski SM, Powell LH, Adler N, Naar-King S, Reynolds KD, Hunter CM, et al. From ideas to efficacy: The ORBIT model for developing behavioral treatments for chronic diseases. Health Psychol. 2015;34(10):971–82. Epub 20150202. doi: 10.1037/hea0000161. PubMed PMID: 25642841; PubMed Central PMCID: PMCPMC4522392.

49. Onken L. Implementation Science at the National Institute on Aging: The Principles of It. Public Policy & Aging Report. 2022;32(1):39–41. doi: 10.1093/ppar/prab034.

50. Onken LS, Carroll KM, Shoham V, Cuthbert BN, Riddle M. Reenvisioning Clinical Science: Unifying the Discipline to Improve the Public Health. Clin Psychol Sci. 2014;2(1):22–34. doi: 10.1177/2167702613497932. PubMed PMID: 25821658; PubMed Central PMCID: PMCPMC4374633.

51. Nuckols TK. Clinical Trial Protocol: A Cluster Randomized Controlled Trial Comparing the Safety Action Feedback and Engagement (SAFE) Loop With an Established Incident Reporting System, ClinicalTrials.gov ID NCT05381441. National Library of Medicine, Clinicaltrials.gov, 2022.

52. Berdahl CT, Henreid AJ, Cohen TN, Coleman BL, Seferian EG, Leang D, et al. Comparing the Safety Action Feedback and Engagement (SAFE) Loop with an established incident reporting system: Study protocol for a pragmatic cluster randomized controlled trial. Contemp Clin Trials Commun. 2023;35:101192. Epub 20230718. doi: 10.1016/j.conctc.2023.101192. PubMed PMID: 37538195; PubMed Central PMCID: PMCPMC10393596.

53. Shojania KG. Incident Reporting Systems: What Will It Take to Make Them Less Frustrating and Achieve Anything Useful? Jt Comm J Qual Patient Saf. 2021;47(12):755–8. Epub 20211012. doi: 10.1016/j.jcjq.2021.10.001. PubMed PMID: 34716115.

54. Goorts K, Dizon J, Milanese S. The effectiveness of implementation strategies for promoting evidence informed interventions in allied healthcare: a systematic review. BMC Health Serv Res. 2021;21(1):241. Epub 20210318. doi: 10.1186/s12913-021-06190-0. PubMed PMID: 33736631; PubMed Central PMCID: PMCPMC7977260.

55. Medves J, Godfrey C, Turner C, Paterson M, Harrison M, MacKenzie L, Durando P. Systematic review of practice guideline dissemination and implementation strategies for healthcare teams and team-based practice. Int J Evid Based Healthc. 2010;8(2):79–89. doi: 10.1111/j.1744-1609.2010.00166.x. PubMed PMID: 20923511.

56. Powell BJ, Waltz TJ, Chinman MJ, Damschroder LJ, Smith JL, Matthieu MM, et al. A refined compilation of implementation strategies: results from the Expert Recommendations for Implementing Change (ERIC) project. Implement Sci. 2015;10:21. Epub 20150212. doi: 10.1186/s13012-015-0209-1. PubMed PMID: 25889199; PubMed Central PMCID: PMCPMC4328074.

57. Last BS, Buttenheim AM, Timon CE, Mitra N, Beidas RS. Systematic review of clinician-directed nudges in healthcare contexts. BMJ Open. 2021;11(7):e048801. Epub 20210712. doi: 10.1136/bmjopen-2021-048801. PubMed PMID: 34253672; PubMed Central PMCID: PMCPMC8276299.

58. Yoong SL, Hall A, Stacey F, Grady A, Sutherland R, Wyse R, et al. Nudge strategies to improve healthcare providers’ implementation of evidence-based guidelines, policies and practices: a systematic review of trials included within Cochrane systematic reviews. Implement Sci. 2020;15(1):50. Epub 20200701. doi: 10.1186/s13012-020-01011-0. PubMed PMID: 32611354; PubMed Central PMCID: PMCPMC7329401.

59. Talat U, Schmidtke KA, Khanal S, Chan A, Turner A, Horne R, et al. A Systematic Review of Nudge Interventions to Optimize Medication Prescribing. Front Pharmacol. 2022;13:798916. Epub 20220125. doi: 10.3389/fphar.2022.798916. PubMed PMID: 35145411; PubMed Central PMCID: PMCPMC8822212.

60. Rogers EM. Diffusion of Innovations, 5th Edition. New York: Free Press; 2003.

61. Greenhalgh T, Robert G, Macfarlane F, Bate P, Kyriakidou O. Diffusion of innovations in service organizations: systematic review and recommendations. Milbank Q. 2004;82(4):581–629. doi: 10.1111/j.0887-378X.2004.00325.x. PubMed Central PMCID: PMC2690184.

62. Moore GC BI. Development of an Instrument to Measure the Perceptions of Adopting an Information Technology Innovation. Institute for Operations Research and the Management Sciences. 1991;2(3):173–239.

63. Valente TW, Davis RL. Accelerating the Diffusion of Innovations Using Opinion Leaders. The Annals of the American Academy of Political and Social Science. 1999;566:55–67.

64. Learn from Defects Tool: Agency for Healthcare Research and Quality, Rockville, MD; 2012. Available from: https://www.ahrq.gov/hai/cusp/toolkit/learn-defects.html.

65. Hartwig SC, Denger SD, Schneider PJ. Severity-indexed, incident report-based medication error-reporting program. Am J Hosp Pharm. 1991;48(12):2611–6. Epub 1991/12/01. PubMed PMID: 1814201.

66. Snyder RA, Abarca J, Meza JL, Rothschild JM, Rizos A, Bates DW. Reliability evaluation of the adapted national coordinating council medication error reporting and prevention (NCC MERP) index. Pharmacoepidemiol Drug Saf. 2007;16(9):1006–13. doi: 10.1002/pds.1423. PubMed PMID: 17523185.

67. Blegen MA, Gearhart S, O’Brien R, Sehgal NL, Alldredge BK. AHRQ’s hospital survey on patient safety culture: psychometric analyses. J Patient Saf. 2009;5(3):139–44. Epub 2009/11/19. doi: 10.1097/PTS.0b013e3181b53f6e. PubMed PMID: 19920453.

68. Sorra JS, Dyer N. Multilevel psychometric properties of the AHRQ hospital survey on patient safety culture. BMC Health Serv Res. 2010;10:199. Epub 2010/07/10. doi: 10.1186/1472-6963-10-199. PubMed PMID: 20615247; PubMed Central PMCID: PMCPmc2912897.

69. DiCuccio MH. The Relationship Between Patient Safety Culture and Patient Outcomes: A Systematic Review. J Patient Saf. 2015;11(3):135–42. Epub 2014/03/04. doi: 10.1097/pts.0000000000000058. PubMed PMID: 24583952.

70. Cohen TN, Nuckols TK, Berdahl CT, Seferian EG, McCleskey SG, Henreid AJ, et al. Training Hospital Nurses to Write Detailed Narratives and Describe Contributing Factors in Incident Reports: The SAFER Education Program. Jt Comm J Qual Patient Saf. 2025;51(4):305–11. Epub 20250110. doi: 10.1016/j.jcjq.2025.01.002. PubMed PMID: 39894711.

71. Haig KM, Sutton S, Whittington J. SBAR: A Shared Mental Model for Improving Communication Between Clinicians. The Joint Commission Journal on Quality and Patient Safety. 2006;32(3):167–75.

72. Carayon P, Hundt AS, Karsh BT, Gurses AP, Alvarado CJ, Smith M, Brennan PF. Work system design for patient safety: the SEIPS model. BMJ Quality & Safety. 2006;15(suppl 1):i50–i8.

73. Shappell SA WD. A Human Error Approach to Aviation Accident Analysis: The Human Factors Analysis and Classification System. Burlington, VT: Ashgate Press; 2003.

74. Classen DC LR, Provost L, Griffin FA, Resar R. Development and evaluation of the Institute for Healthcare Improvement Global Trigger Tool. Journal of Patient Safety. 2008;4(3):169–77.

75. SOPS Hospital Survey: Agency for Healthcare Research and Quality, Rockville, MD; 2022. Available from: https://www.ahrq.gov/sops/surveys/hospital/index.html.

76. Hempel S, Shekelle PG, Liu JL, Sherwood Danz M, Foy R, Lim Y-W, et al. Development of the Quality Improvement Minimum Quality Criteria Set (QI-MQCS): a tool for critical appraisal of quality improvement intervention publications. BMJ Quality && Safety. 2015;24(12):796. doi: 10.1136/bmjqs-2014-003151.

77. Taylor SL, Dy S, Foy R, Hempel S, McDonald KM, Ovretveit J, et al. What context features might be important determinants of the effectiveness of patient safety practice interventions? BMJ Qual Saf. 2011;20(7):611–7. Epub 20110526. doi: 10.1136/bmjqs.2010.049379. PubMed PMID: 21617166.

78. Ogrinc G, Davies L, Goodman D, Batalden P, Davidoff F, Stevens D. SQUIRE 2.0 (Standards for QUality Improvement Reporting Excellence): revised publication guidelines from a detailed consensus process. BMJ Qual Saf. 2016;25(12):986–92. Epub 20150914. doi: 10.1136/bmjqs-2015-004411. PubMed PMID: 26369893; PubMed Central PMCID: PMCPMC5256233.

79. Damschroder LJ, Reardon CM, Widerquist MAO, Lowery J. The updated Consolidated Framework for Implementation Research based on user feedback. Implementation Science. 2022;17(1):75. doi: 10.1186/s13012-022-01245-0.

80. Kistin C, Silverstein M. Pilot Studies: A Critical but Potentially Misused Component of Interventional Research. Jama. 2015;314(15):1561–2. doi: 10.1001/jama.2015.10962. PubMed PMID: 26501530; PubMed Central PMCID: PMCPMC4917389.

81. Parmelli E, Flodgren G, Fraser SG, Williams N, Rubin G, Eccles MP. Interventions to increase clinical incident reporting in health care. Cochrane Database Syst Rev. 2012;2012(8):Cd005609. Epub 20120815. doi: 10.1002/14651858.CD005609.pub2. PubMed PMID: 22895951; PubMed Central PMCID: PMCPMC4171121.

