## Appendix survey for "The Safety Action Feedback and Engagement (SAFE) Loop: Initial Testing and Refinement of a Novel Intervention to Enhance Hospital Incident Reporting and Patient Safety"

**Measures Derived from Survey**

| | Type | Scale | Timing |
|---|---|---|---|
| Confidence in Reporting | Composite measure, novel | Percent positive response (0-100%) | All |
| Communication About Error | Composite measure, AHRQ SOPS | Percent positive response (0-100%) | All |
| Reporting Patient Safety Events | Composite measure, AHRQ SOPS | Percent positive response (0-100%) | All |
| Blame-free Culture | Individual measure, AHRQ SOPS | Percent positive response (0-100%) | All |
| Incident Report Submissions | Individual measure, AHRQ SOPS | 0-4+, Median, IQR | All |
| Time per Report | Individual measure, novel | Continuous (0-90) | All |

**Survey Instrument**

| Item Stem | Response Options | Source |
|---|---|---|
| *How often do the following things happen in your unit?* | | |
| 1. Nurses are informed about errors that happen in this unit. | Response Options:<br>1 – Never<br>2 – Rarely<br>3 – Sometimes<br>4 – Most of the time<br>5 – Always<br>6 -- Does not apply or don't know | AHRQ SOPS Hospital Survey 2.0, topic 6, communication about error, Q C1. |
| 2. When errors happen in this unit, nurses discuss ways to prevent them from happening again. | | AHRQ SOPS Hospital Survey 2.0, topic 6, communication about error, Q C2. |
| 3. In this unit, nurses are informed about changes that are made based on incident reports. | | AHRQ SOPS Hospital Survey 2.0, topic 6, communication about error, Q C3. |
| 4. When staff make errors, this unit focuses on learning rather than blaming individuals. | | AHRQ SOPS Hospital Survey 2.0, topic 4, response to error, Q A10. |
| 5. When a mistake is caught and corrected before reaching the patient, how often is this reported using the incident reporting system? | | AHRQ SOPS Hospital Survey 2.0, topic 8, reporting patient safety events, Q D1. |
| 6. When a mistake reaches the patient and could have harmed the patient, but did not, how often is this reported using the incident reporting system? | | AHRQ SOPS Hospital Survey 2.0, topic 8, reporting patient safety events, Q D2. |
| *How much do you agree or disagree with the following statement about your unit?* | | |
| 7. How confident are you in knowing when to complete an incident report for a medication safety event? | Consider the following questions related to completing an incident report for a medication safety event.<br>Response Options:<br>1 – Not at all confident<br>2 – Slightly confident<br>3 – Moderately confident<br>4 – Very confident<br>5 – Extremely confident | Novel |
| 8. How confident are you in knowing what information to include when completing an incident report for a medication safety event? | | Novel |

| | | |
|---|---|---|
| 9. In the last 6 months, how many medication safety events have you personally reported via the incident reporting system? | 1 – None<br>2 – 1<br>3 – 2<br>4 – 3<br>5 – 4 or more | AHRQ SOPS Hospital Survey 2.0, topic [11], number of events reported, Q D3. |
| [If response to item above was 2 through 5]<br>10. Please think of the last time you completed an incident report for a medication safety event. How many minutes did it take you to complete the report? | [Response options in increments of 5 min, e.g., 5, 10, 15, up to 90…) | Novel |

**Coding of Measures for Analysis:**

*Composite measure "Confidence in Reporting:"* Use methods below for calculating composite measures. Positive responses: very confident or extremely confident.

- How confident are you in knowing when to complete an incident report for a medication safety event?
- How confident are you in knowing what information to include when completing an incident report for a medication safety event?

*Composite measure "Communication About Error:"* The extent to which staff are informed when errors occur, discuss ways to prevent errors, and are informed when changes are made. Positive responses: most of the time or always. (Q C1-C3)

- Nurses are informed about errors that happen in this unit.
- When errors happen in this unit, nurses discuss ways to prevent them from happening again.
- In this unit, nurses are informed about changes that are made based on incident reports.

*Composite measure "Reporting Patient Safety Events:"* The extent to which mistakes of the following types are reported: (1) mistakes caught and corrected before reaching the patient and (2) mistakes that could have harmed the patient but did not. (Q D1-2)

- When a mistake is caught and corrected before reaching the patient, how often is this reported using the incident reporting system?
- When a mistake reaches the patient and could have harmed the patient, but did not, how often is this reported using the incident reporting system?

*Individual measure "Blame-free Culture:"* When staff make errors, this unit focuses on learning rather than blaming individuals. Positive responses: most of the time or always.

*Individual measure "Incident Report Submissions:"* In the last 6 months, how many medication safety events have you personally reported via the incident reporting system?

*Individual measure "Time per Report:"* Please think of the last time you completed an incident report for a medication safety event. How many minutes did it take you to complete the report?

Methods for calculating percent positive response and, if applicable, creating a composite measure:

- Identify respondents to the survey overall. Exclude non-respondents.
- For an individual item of interest,
    - Identify and count responses. Exclude those with missing responses and does not apply/do not know responses to that item.
    - Identify and count positive responses.

    - Calculate the percent positive response as the count of positive responses divided by the total count of responses.
- To calculate the composite measure, average the percent positive scores for each item included in the composite measure.
