## Supplementary material for "The Safety Action Feedback and Engagement (SAFE) Loop: Initial Testing and Refinement of a Novel Intervention to Enhance Hospital Incident Reporting and Patient Safety": IRB approval letter

Cedars Sinai

Teryl K. Nuckols, MD, MSHS
Division of General Internal Medicine
Department of Medicine
Cedars-Sinai Medical Center
8700 Beverly Blvd
Los Angeles, CA 90048

June 2, 2025

Dear Dr. Nuckols,

This letter is to verify that your research project, the Safety Action Feedback and Engagement (SAFE) Loop, was reviewed and approved by the Cedars-Sinai Institutional Review Board (IRB). Two IRB records are relevant to this project.

**Pro00055431: Safety Action Feedback and Engagement (SAFE) Loop**
On January 2, 2019, the IRB reviewed this project and determined that it did not meet the FDA and DHHS definition of human subjects' research, meaning that it is not regulated. The review was last updated on June 26, 2021.

**STUDY00001025: Safety Action Feedback and Engagement (SAFE) Loop**
The IRB approved this project under expedited review categories 5 and 7. The approval has been active since May 18, 2021 and remains active today. The IRB approval does not have an expiration date, and the study will remain approved until the investigators choose to close it.

Sincerely,
Emily

Emily Moody

Digitally signed by Emily Moody
Date: 2025.06.02 17:11:41 -07'00'

Emily Moody, CIP
IRB Analyst II, Office of Research Compliance & Quality Improvement
